# Personalized Synergy-based Functional Electrical Stimulation Improves Lower Limb Motor Functions of Chronic Stroke Survivors by Restoring Gait Control Modules

**DOI:** 10.1101/2025.04.30.25326772

**Authors:** Xiaoyu Guo, Kelvin Y.S. Lau, Minglei Bai, Richard Liu, Borong He, Jodie J. Xie, Jiayin Lin, Vivian K.H. Yuen, Peter P.K. Chan, Summer L.S. Law, Junhao Wang, Sin Ming Li, Chih-Hong Chou, Chao-ying Chen, Gladys L.Y. Cheing, Patrick W.H. Kwong, Ning Lan, Roy T.H. Cheung, Rosa H.M. Chan, Vincent C.K. Cheung

**Affiliations:** Department of Electrical Engineering, City University of Hong Kong, Hong Kong; School of Biomedical Sciences, Gerald Choa Neuroscience Institute, and KIZ-CUHK Joint Laboratory of Bioresources and Molecular Research of Common Diseases, The Chinese University of Hong Kong, Hong Kong; Department of Rehabilitation Sciences, The Hong Kong Polytechnic University, Hong Kong; Li Ka Shing Faculty of Medicine, University of Hong Kong, Hong Kong; Physiotherapy Department, Tseung Kwan O Hospital, Hong Kong; School of Biomedical Engineering and Med-X Research Institute, Shanghai Jiao Tong University, Shanghai, China; School of Physical Therapy and Graduate Institute of Rehabilitation Science, College of Medicine, Chang Gung University, Taoyuan, Taiwan; Department of Physical Medicine and Rehabilitation, Linkou Chang Gung Memorial Hospital, Taoyuan, Taiwan; Richard and Loan Hill Department of Biomedical Engineering, University of Illinois at Chicago, Chicago, USA; School of Health Sciences, Western Sydney University, Sydney, NSW, Australia

**Keywords:** Stroke, Rehabilitation, FES, muscle synergy

## Abstract

Conventional motor rehabilitation for stroke remains labor intensive and shows limited efficacy for chronic survivors. Non-invasive functional electrical stimulation (FES) of the neuromuscular system is promising for restoring mobility. However, traditional FES interventions for stroke are limited by single-channel protocols that target isolated muscles and lack integration with neuromotor control strategies, thus failing to address the heterogeneity of post-stroke motor deficits. To overcome these challenges, we employed a personalized FES paradigm grounded in muscle synergies — neuromotor modules that coordinate multimuscle activation during movement. By leveraging muscle synergies as biomarkers of motor impairment, our intervention delivers coordinated multi-muscle stimulation that mimics the healthy muscle patterns absent in each stroke survivor. Compared with sham group (***N* = 10**), chronic stroke survivors in the treatment group (***N* = 23**) demonstrated higher synergy similarity to the normative synergies after treatment (***p* = 0.01**). Notably, this improvement correlated with the gain in lower-limb Fugl-Meyer (FM) score (**△**FM **= 2.6** vs. **0.7**, ***p* = 0.04**) and enhancement in gait symmetry. Our study shows, for the first time, that muscle synergy-based FES can address the one-size-fits-all limitation of conventional FES by offering a personalized, efficacious, and neuroscience-based intervention that may improve gait kinematics and motor control even in chronic stroke survivors through restoration of muscle synergies.

## 1 Introduction

Stroke is the third leading global cause of mortality and disability, imposing significant socioeconomic burdens and healthcare costs [1]. Over 60% of stroke survivors suffer from walking impairments, which severely limits independence and quality of life [2]. Therefore, restoring the ability to walk independently is one of the central goals of poststroke motor rehabilitation [3]. Conventional physiotherapies such as repetitive gait training rely heavily on physical therapists to manually guide patients through high-intensity exercises [4]. While effective, these approaches are labor-intensive and often fail to meet the high demands of sustained, repetitive training from a large number of stroke survivors, particularly given the global shortages of rehabilitation specialists [5]. Other widely used interventions such as Bobath therapy and robotic assistance demonstrate efficacy primarily in acute and subacute stroke patients, thus leaving chronic survivors with limited rehabilitative options [4, 6]. Addressing these limitations of existing rehabilitative interventions requires innovative strategies to maximize gait recovery across all stages of impairment [7].

Functional electrical stimulation (FES), a rehabilitation intervention that applies controlled electrical currents to the neuromuscular system [8], has emerged as a promising tool for restoring motor functions in neurological conditions such as spinal cord injury [9, 10], cerebral palsy [11], and stroke [12–14]. Clinically, FES that stimulates the neuromuscular system through surface electrodes has been used for decades to enhance post-stroke lower limb mobility by strengthening ankle dorsiflexors, improving muscle coordination, and reducing foot-dragging [15–18]. Its non-invasive nature and adaptability make it particularly appealing for stroke rehabilitation for which individualized treatment plans are critical.

Despite its potential, traditional FES applications face significant limitations. Current protocols predominantly rely on single-channel stimulation (e.g., one that targets the tibialis anterior) [19–21], which enhances activation of a specific muscle or muscle group but yields only marginal improvements in walking ability [22]. While this approach can reinforce the activation of an individual muscle such as any ankle dorsiflexor, it often results in only basic improvement in locomotor outcome. The gait patterns observed following FES-based gait retraining may remain rigid, potentially affecting gait efficiency despite improvements in overall function. Furthermore, traditional FES demands prolonged, resource-intensive interventions. A meta-review study [23] found that FES interventions for the lower limb typically range from 12 to 48 sessions in previous studies. For example, protocols can include 30 sessions [24], 20 sessions [21], or 60 sessions [20].

More critically, current FES rehabilitation approaches often lack integration with a neuromotor control strategy, limiting their ability to adapt stimulation timing, intensity, or dosage to a patient’s residual motor capacity [25–27]. They operate without regard to any neurophysiological markers that may reflect the specific motor deficit of the stroke survivor. This one-size-fits-all approach overlooks the immense heterogeneity of post-stroke motor impairments. Probably because of this reason, these traditional training methods demand significant time and human resources to achieve just limited efficacy on average. Here, we argue that FES strategies based on neuromotor control principles could bridge this gap, enabling tailored interventions for stroke survivors with diverse residual motor functions of stroke survivors [28]. Such personalized interventions promise to achieve more naturalistic and sustainable improvements in walking ability, particularly for chronic stroke survivors with complex rehabilitation needs.

To address these limitations of traditional FES for stroke rehabilitation, we pursue here a relatively novel FES paradigm based on the muscle synergy principle. Muscle synergies are hypothesized as neural modules of motor control that coordinate the spatial activation patterns of multiple muscles together for movement execution [29, 30]. Derived from multi-muscle electromyography (EMG) signals via factorization algorithms [31], muscle synergies refer to quantitative estimates of muscle coordination patterns [32] encoded by spinal interneurons, but modulated in their temporal recruitment by descending commands from the higher motor areas and afferent sensory feedback [33, 34]. Furthermore, synergies may serve as biomarkers that indicate the motor deficits of stroke survivors with cortical lesions [35], especially when the synergies exhibit distinct patterns such as merging and fractionation of muscle synergies [36]. Importantly, the use of synergy patterns as quantifiable biomarkers of motor impairment offers the possibility of designing personalized intervention tailored to each patient’s residual neuromuscular capacity.

Our FES intervention here leverages these insights to deliver coordinated, multimuscle stimulation that mimics natural neuromuscular coordination. Unlike conventional single-channel FES which reinforces a single isolated muscle and risks unnatural gait adaptations, our approach reinforces functional muscle groups in spatiotemporal patterns that resemble the normative muscle synergies. Specifically, for each stroke survivor, we identified the normative synergies that were missing in the survivor’s affected leg, and constructed an FES waveform by summing these absent normative synergies that scaled temporally by their expected, normative profiles. We hypothesize that this strategy, by providing proprioceptive and afferent feedback through the designed FES stimulus, may trigger a specific neuroplastic mechanism that guides reconfiguration of the residual neural circuits to produce normalized motor patterns through the combination of the normative muscle synergies. By recruiting and reinforcing normal synergies through neuroplasticity, this approach can promote more natural and functional gait recovery in diverse stroke survivors with wide-ranging motor deficits.

We evaluated this paradigm in a cohort of chronic stroke survivors, comparing a treatment group (*N* = 23) receiving synergy-based FES with a control group (*N* = 10) receiving sham stimulation. Assessments including lower-limb Fugl-Meyer (FM) score, gait kinematics, and muscle synergies were conducted before, immediately after, and one month after the completion of intervention. After FES training, the treatment group demonstrated significantly greater improvements in FM score, and exhibited muscle synergies that become more similar to the normative synergies selected for FES construction. Importantly, such increase in synergy similarity was accompanied by more naturalistic gait kinematics and improved step symmetry. These findings argue that synergy-based FES is a viable strategy of personalized post-stroke motor rehabilitation that can lead to motor functional improvement by promoting the recruitment of closer-to-normal muscle synergies, thereby resulting in more naturalistic movement after FES.

## 2 Results

### 2.1 Strategy of personalizing FES rehabilitative intervention based on muscle synergies

For each stroke patient, personalized FES pattern was constructed using muscle synergies from a matched healthy subject that were dissimilar to the patient’s stroke-affected synergies (scalar product *<* 0.8). For two patients with only one normative synergy with similarity below 0.8, the scalar product threshold was adjusted to 0.85. These selected normative synergies were absent on the patient’s stroke-affected leg; the FES pattern therefore targets the deficient motor modules by integrating their expected normative synergistic muscle pattern and temporal activation profiles into the FES design to guide neuromuscular retraining (Fig. 1). We hypothesized that in stroke survivors, stimulation of these prioritized synergies recalibrates the residual neural circuits through sensory feedback mechanisms towards the normative muscle patterns, thereby promoting functional recovery. To evaluate this hypothesis, we conducted longitudinal comparisons in both FES and sham groups, measuring changes in FM scores, joint kinematics, and muscle synergy similarity.

**Fig. 1.**
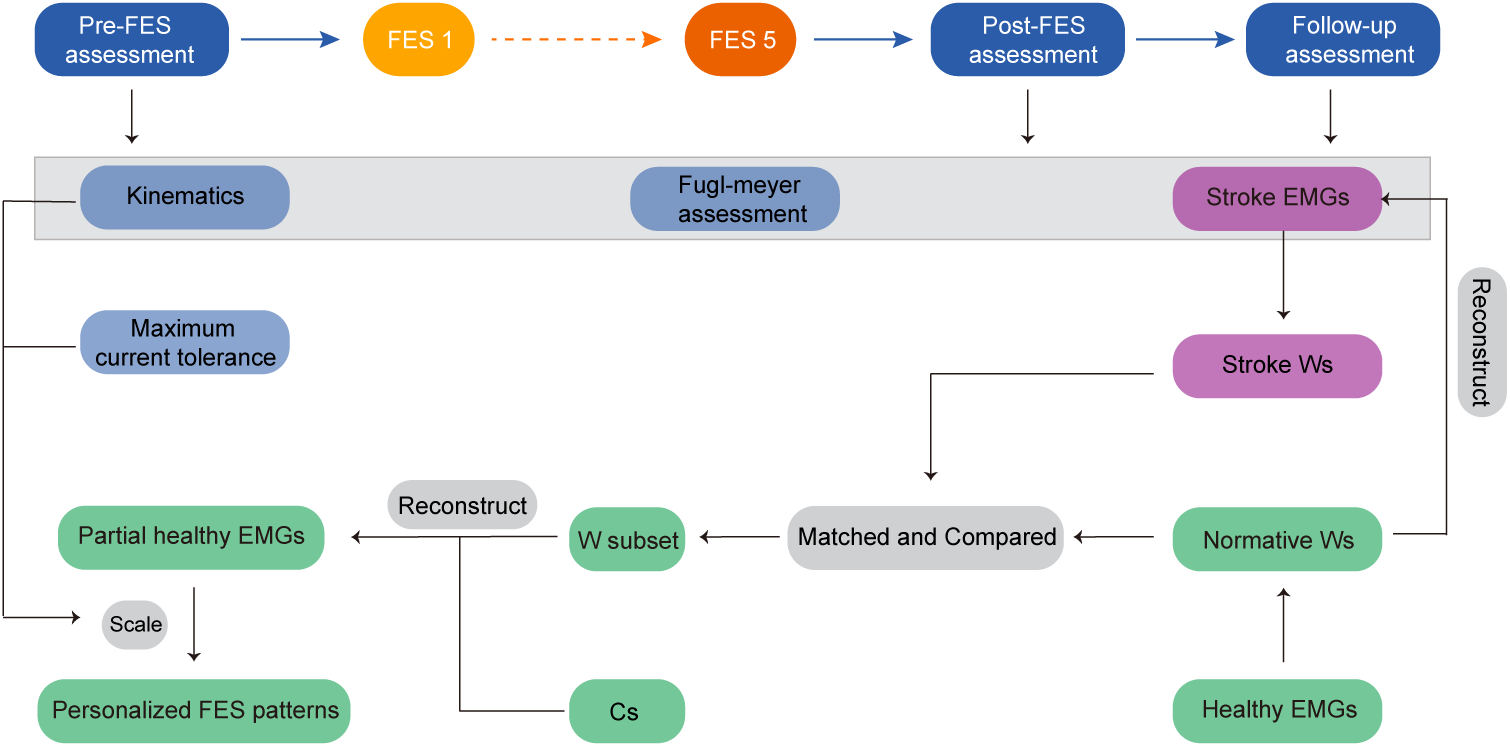
The framework for personalized synergy-based FES pattern design for lower-limb rehabilitation in chronic stroke survivors. The protocol begins with pre-FES assessments including FM assessment, gait kinematics, EMG recordings, and maximum current tolerance testing of each muscle. Five FES sessions were then conducted for stroke survivors over 20 *±* 11 days. Post-FES and follow-up assessments were conducted immediately and one month (*>* 30 days) after the completion of intervention. For FES waveform construction, normative synergies of the selected normal subject were paired with stroke-affected synergies via scalar product similarity. The normative synergies with a similarity below 0.8 were selected and multiplied by their corresponding temporal coefficients to reconstruct the partial normative EMG signals. These partial EMG patterns were temporally rescaled to align with the stance ratio of each stroke survivor during natural walking. The stimulation amplitude for each muscle was adjusted to the maximum current tolerance tested in pre-assessment.

### 2.2 Synergy-based FES improves Fugl-Meyer scores

Before intervention, the attributes of the FES and sham groups were statistically homogeneous. No significant differences were found in stroke chronicity (two-tailed unpaired t-test, *p* = 0.33), lesion type (Fisher’s exact test, *p* = 1.0), age (two-tailed unpaired t-test, *p* = 0.14) or gender (Fisher’s exact test, *p* = 1.0).

Longitudinal analysis of lower-extremity FM scores (maximum = 34) revealed significant post-training improvements in the FES group. At the pre-FES stage, both groups exhibited comparable FM scores (FES group: 21.4 ± 5.4; sham group: 21.7 ± 6.5, two-tailed unpaired t-test, *p* = 0.9). At the post-FES stage, FES group showed significantly higher FM scores (24 ± 4.5 vs. pre-FES, *p* = 2.2 × 10*^−^*^5^, two-tailed paired t-test) while the sham group demonstrated minimal change (22.4 ± 5.6 vs. baseline, *p >* 0.05) (Fig. 2a). At one-month follow-up, this improvement persisted in the FES group (24±5.4, *p* = 1.6×10*^−^*^4^ vs. baseline) while the sham group remained unchanged (22.4 ± 5.8). Quantifications of FM-score changes across sessions further validated these findings (Fig. 2e). The FES group achieved an average improvement of 2.6 points post-FES, increasing to 2.9 points at follow-up with 50% of the participants exceeding 3 points of change. The sham group, however, showed minor gains (0.7 points at both post-FES and follow-up). The effect size of the FES group was large (*Cohen^′^sd* = 1.12). This inter-group difference highlights the sustained neuroplastic benefits from coordinated, synergy-driven stimulation.

**Fig. 2.**
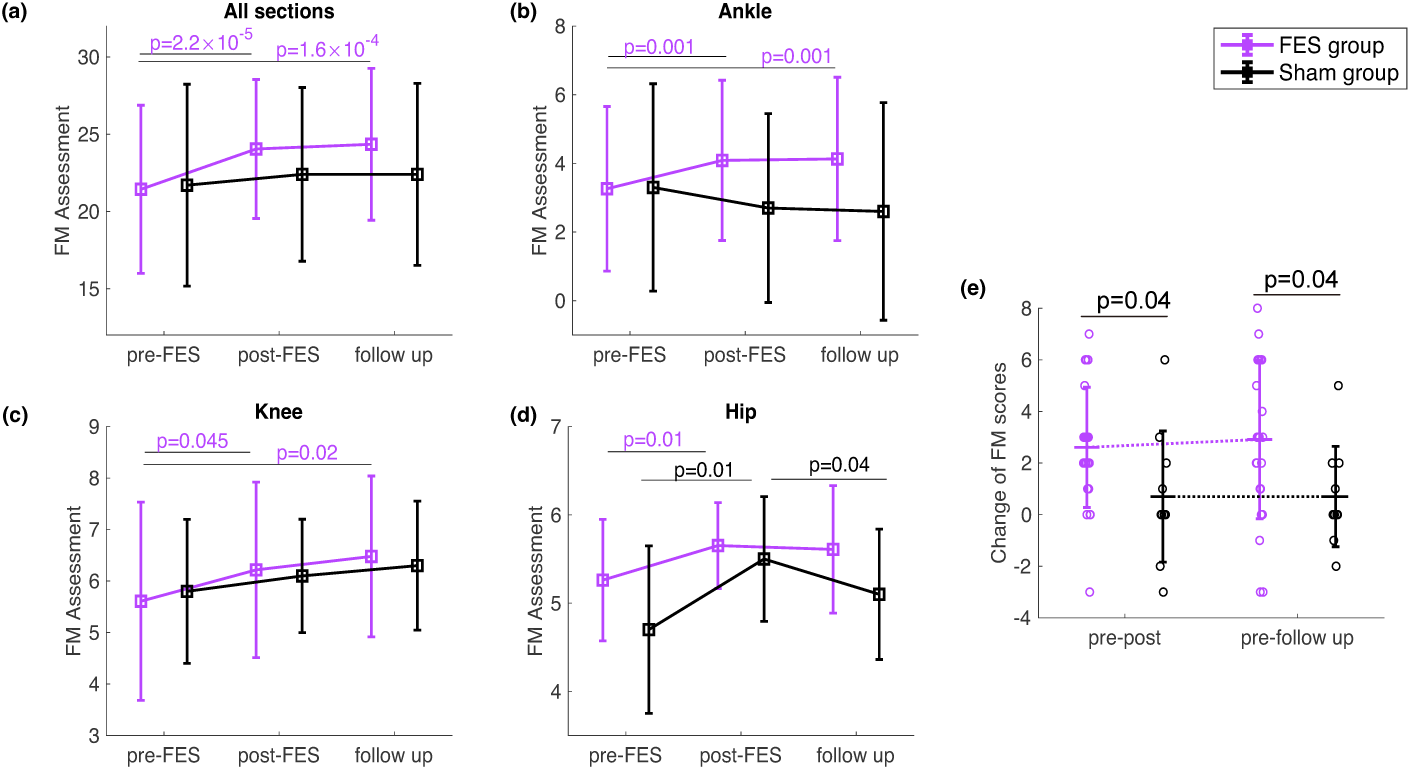
Fugl-Meyer (FM) assessments across intervention stages. Mean *±* standard deviation FM scores for the FES group (*N* = 23, purple) and sham group (*N* = 10, black) at pre-FES, post-FES, and one-month follow-up. Two-tailed unpaired t-tests compared group differences. (a) Total FM scores (max 34). (b) Ankle subsection scores: dorsiflexion of flexor/extensor synergy, plantarflexion of seated/standing tasks (max 8). (c) Knee subsection scores: flexion of flexor/extensor synergy and seated/standing tasks (max 8). (d) Hip subsection scores: flexion, extension, and adduction (max 6). (e) Longitudinal changes: Δ post- vs. pre-FES and Δ one-month vs. pre-FES.

More specifically, analyses of the ankle, knee, and hip sub-scores of the FM assessment corroborate the above trends (Fig. 2b–d). The FES group exhibited higher FM sub-scores for all three joints at post-FES (*p <* 0.05), and these FM sub-scores remained non-decreasing at the follow-up stage. In contrast, the sham group showed no statistical increase except for the hip sub-score (*p* = 0.01), but it decreased back to the initial level at follow-up. These findings suggest that synergy-based FES enhances multi-joint motor recovery, especially in ankle- and knee-related motor functions.

### 2.3 Synergy-based FES produces more naturalistic kinematics on the affected side

To account for the heterogeneity in the changes in gait kinematics, we have found it helpful to stratify participants in the FES group into subgroups based on stroke chronicity (range: 8–232 months) when evaluating the pre-to-post kinematic changes. Patients were divided into three cohorts: short-term chronicity (8–23 months; 25th percentile, *N* = 8), mid-term chronicity (40 − 75 months, *N* = 8), and long-term chronicity (87–232 months; 75th percentile, *N* = 7). Sagittal-plane joint angles (ankle, knee, hip) were averaged within each subgroup and aligned temporally from heel strike to heel strike for pre- and post-FES comparisons (Fig. 3).

**Fig. 3.**
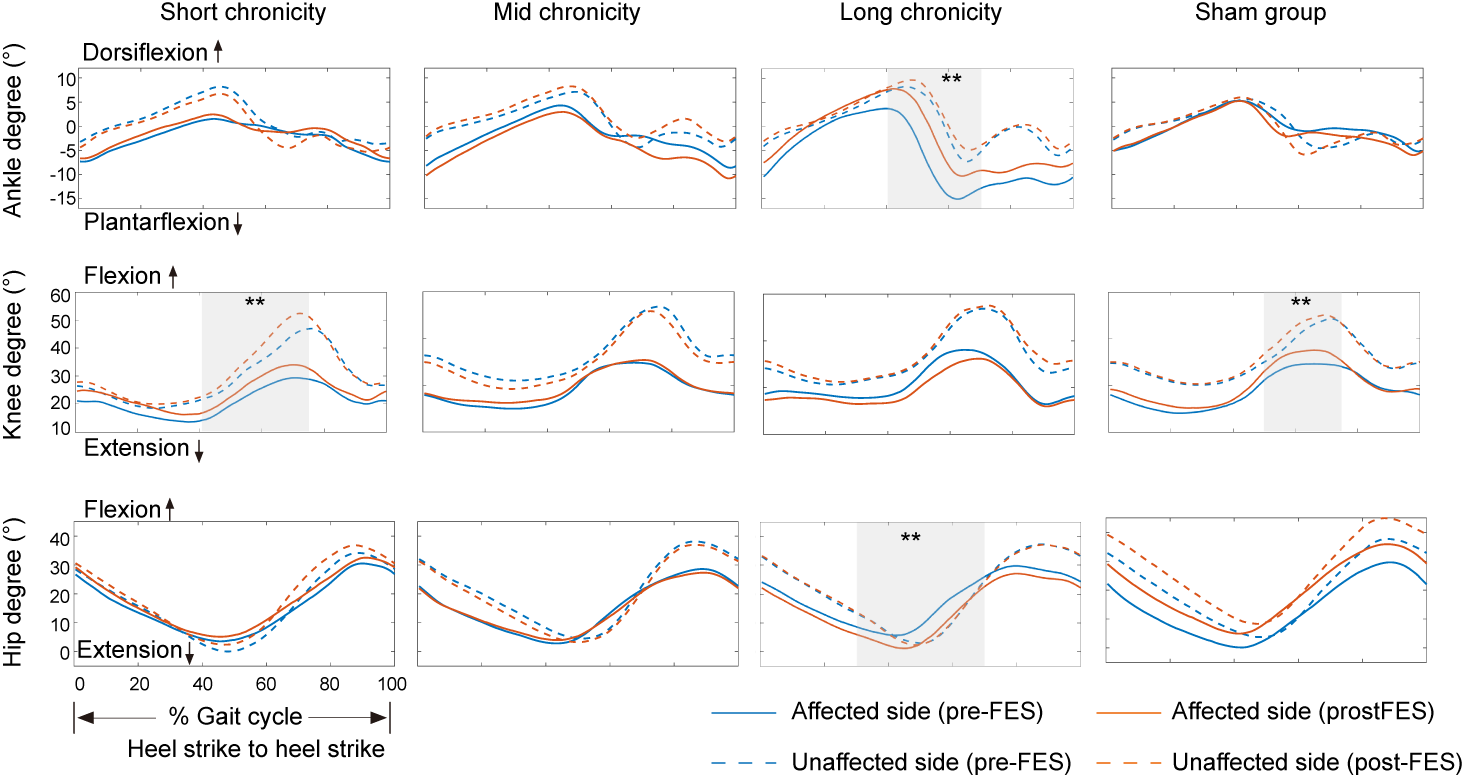
Sagittal plane joint angles before and after FES intervention. The FES group was separated into three subgroups, short-term (8–23 months; *N* = 8), mid-term (40–75 months, *N* = 8), and longterm chronicity (87–232 months; *N* = 7), according to the post-stroke chronicity. In each subfigure, the joint angle is plotted in a completed gait cycle from heel strike to heel strike. The affected sides are in solid lines and the unaffected sides are in dashed lines. The pre-FES and post-FES are illustrated in blue and orange colors respectively. The significant difference (*p <* 0.05, Two-tailed unpaired t- tests) of affected before and after the interventions is shown in the grey shadow with black stars.

Kinematic improvements varied by stroke chronicity. As illustrated in Fig. 3, the short-term subgroup exhibited bilateral increase in late-stance knee flexion after intervention (*p <* 0.05; shaded regions) while no significant changes were observed in the mid-term subgroup. Notably, in the long-term subgroup, kinematics of the affected leg demonstrated improved ankle dorsiflexion and hip extension during late stance (*p <* 0.05), approximating unaffected-side kinematics. The sham group showed increases in affected-side knee flexion (*p <* 0.05) but with bilateral hip angle exhibited a consistent upward shift, which may suggest compensatory adaptations rather than targeted recovery.

To quantify kinematic improvements, we analyzed the across-session changes in peak ankle dorsiflexion, knee flexion, and hip extension at the stance phase across groups (Fig. 4a, b and c). The long-term FES subgroup again showed the greatest improvement in ankle dorsiflexion (*p* = 0.01 vs. sham). Although hip extension gains were also the largest in the long-term FES subgroup, intergroup differences were not statistically significant. Beyond discrete metrics, we assessed the alignment of ankle- and hip-angle trajectories of the stroke patients with the normative angle patterns of the healthy controls matched to the patients for FES construction. The mid- and long-term subgroups showed higher improvement in ankle-angle alignment than the short-term and sham groups even though inter-group differences were not significant (Fig. 4d). Notably, the long-term subgroup showed the greatest alignment improvement in hip angle, significantly exceeding the short-term subgroup (*p* = 0.04), mid-term subgroup (*p* = 0.02) and sham group (*p* = 0.05, Fig. 4e). Vertical center-of-mass (COM) excursions were further analyzed to evaluate gait stability (Fig. 4f). The long-and short-term FES subgroups exhibited significantly reduced COM variance post- FES (*p <* 0.05 vs. sham), reflecting improved step-to-step stability during overground walking.

**Fig. 4.**
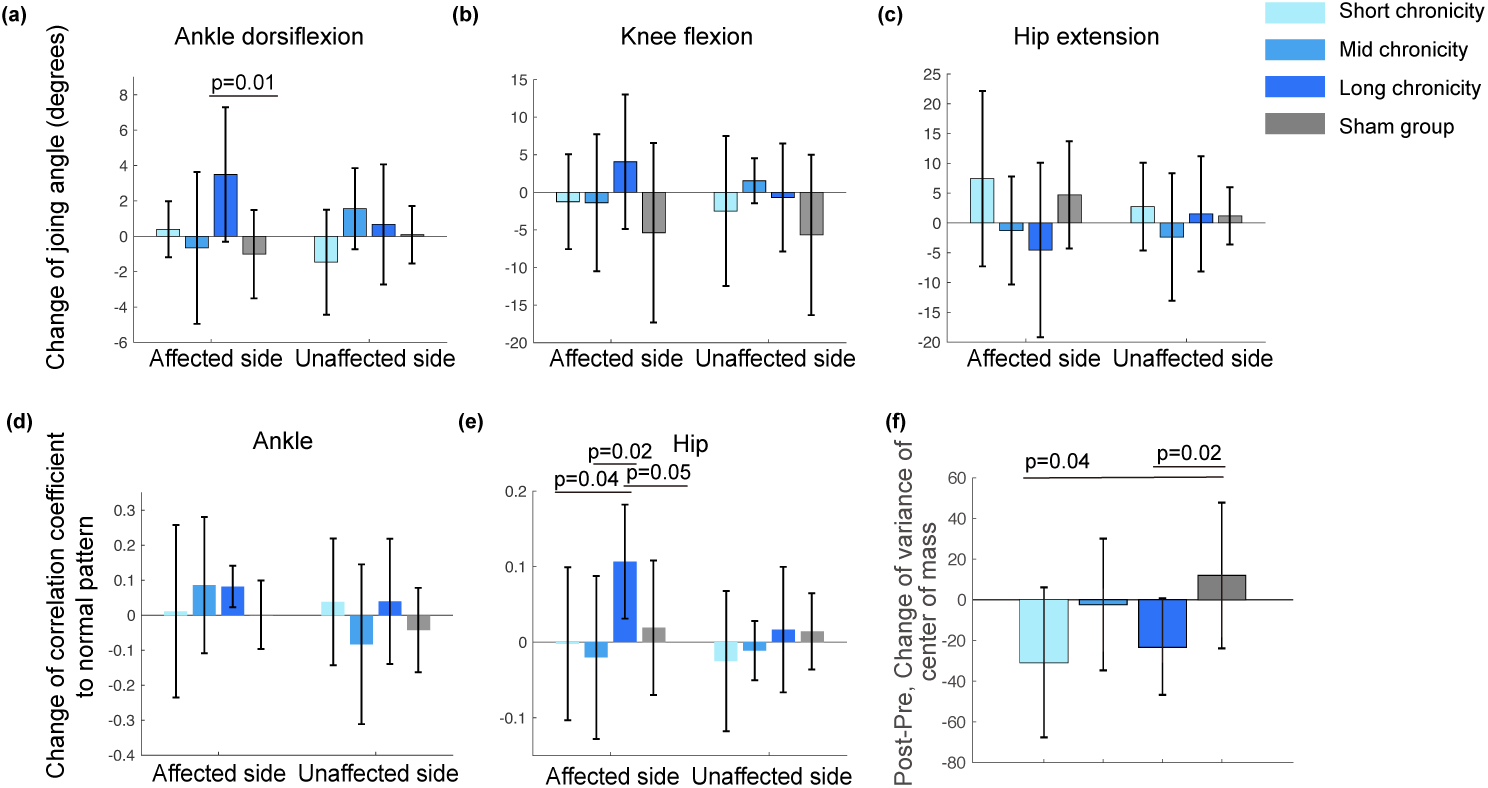
Changes in joint kinematics and center-of-mass stability between pre- and post-FES for the FES group (stratified by short/mid/long chronicity) compared to the sham group. (a–c) Changes in peak joint angles during late stance: ankle dorsiflexion (a), knee flexion (b), and hip extension (c), represented by boxplots (mean *±* standard deviation). (d–e) Correlation coefficient changes between post-FES joint angles (ankle (d); hip (e)) and normative sagittal-plane patterns. (f) Changes of center-of-mass variance post-FES. Two-tailed unpaired t-tests

### 2.4 Synergy-based FES modifies stroke-affected synergies to be more similar to normative synergies

Synergy-based FES selectively enhanced the similarity between the stroke-affected synergies selected for FES intervention and their corresponding normative synergies used for FES construction. Before FES, the FES and sham groups exhibited comparable scalar product similarity between the selected affected-side synergies and the normative synergies (FES group: 0.57 ± 0.24; sham group: 0.66 ± 0.13), but the FES group exhibited increased similarity to the normative synergies (0.71 ± 0.23 vs. baseline, *p* = 4.8 × 10*^−^*^4^)(Fig. 5a) at post-FES stage, with this increase sustained at the follow-up session. This post-FES improvement in the FES group (△ = 0.13 ± 0.28) significantly exceeded the changes in the sham group (△ = 0.02 ± 0.19, *p* = 0.01) (Fig. 5c). On the other hand, the non-targeted affected-side synergies exhibited reduced similarity to the normative synergies in both the FES and sham groups at post-FES (Fig. 5b). When the entire synergy set of each subject was considered in the similar- ity calculations, both the scalar product (Fig. 5d) and subspace similarity (Fig. 5e) of the whole synergy set to the normative remained stable across all sessions in the FES group, but the sham group showed some small fluctuations. These results indicate that synergy-based FES selectively recalibrates the impaired synergies towards their targeted synergies while the non-targeted synergies may exhibit non-specific disruptions. Figure 6 illustrates the modification of stroke-affected muscle synergies in a representative patient before and after the intervention, including their alignment with the normative synergies and the resulting similarity quantification.

**Fig. 5.**
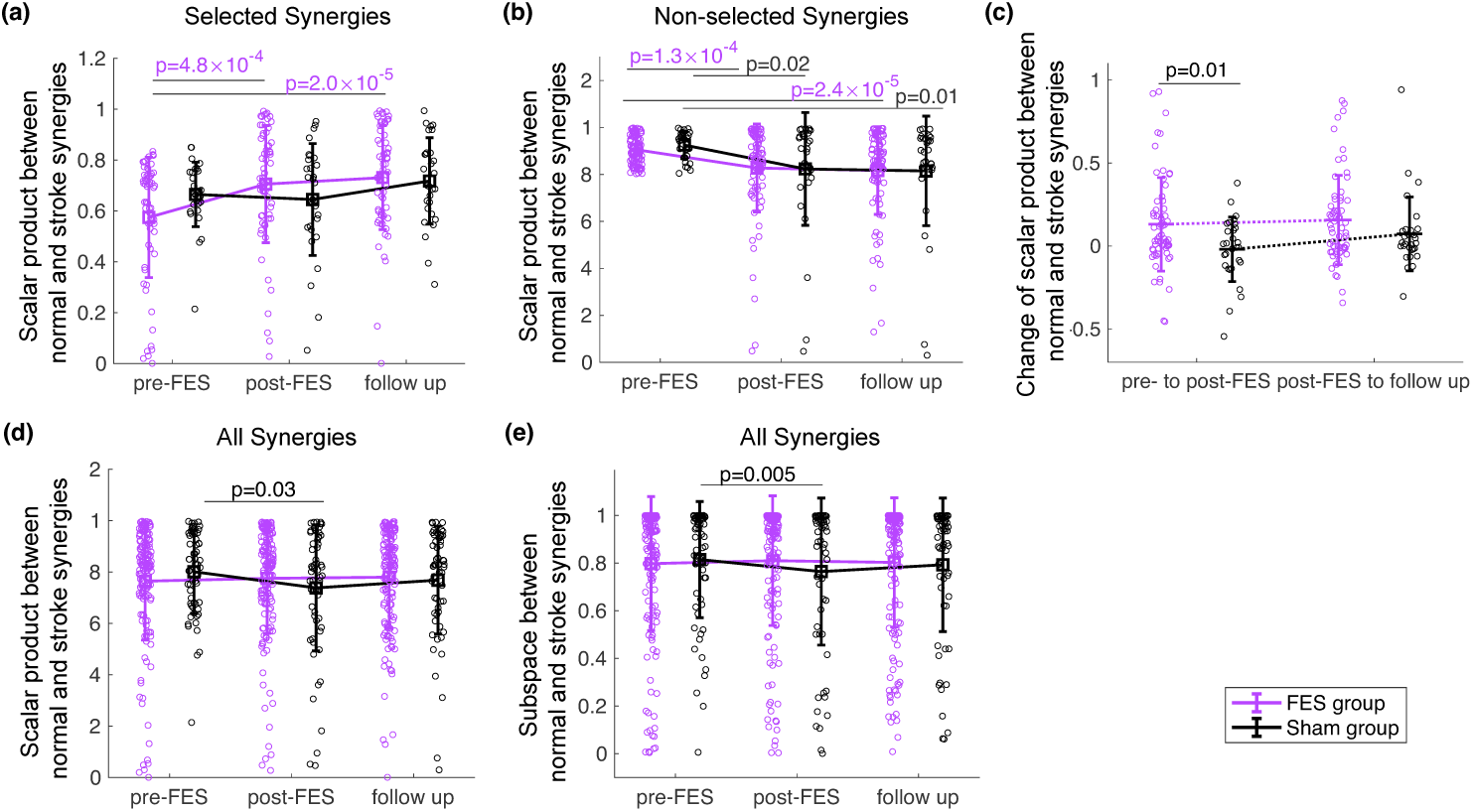
Similarity of muscle synergies across pre-FES, post-FES, and follow-up stages. Two-tailed unpaired t-tests assessed group differences. (a) Selected synergies (targeted for intervention). (b) Nonselected synergies (not targeted). (c) Δ similarity (post vs. pre) and Δ similarity (follow-up vs. post) for selected synergies. (d) Similarity trends for all synergies (selected + non-selected). (e) Subspace similarity between stroke and normative synergy sets.

**Fig. 6.**
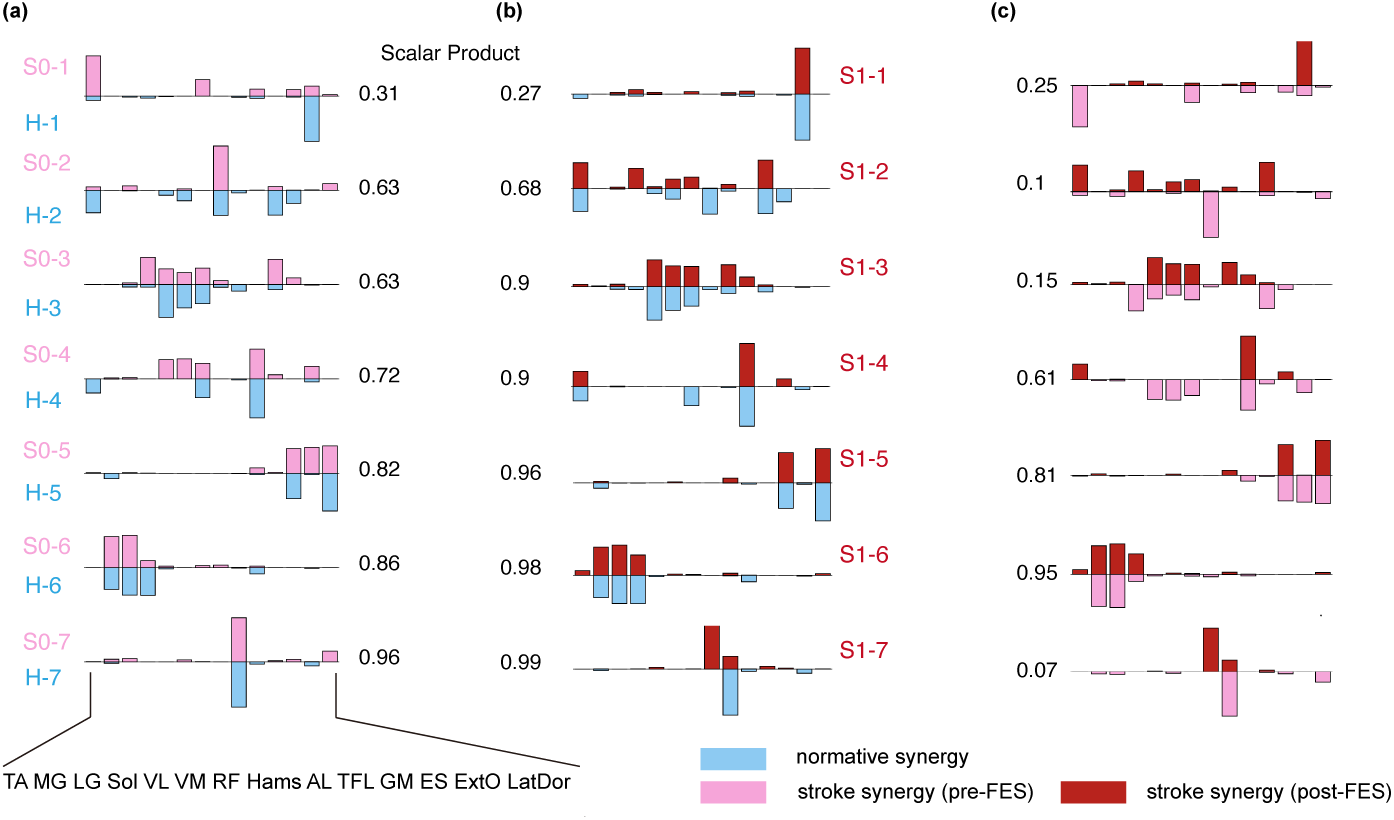
The longitudinal changes in stroke-affected muscle synergies and their alignment with normative synergies. (a) matches pre-FES stroke synergies (pink) to normative synergies (blue), with scalar product similarity scores indicated. (b) demonstrates post-FES stroke synergies (darker red) realigned with normative synergies, highlighting improved overlap. To directly visualize interventioninduced modifications, (c) contrasts pre- and post-FES stroke synergies to show modifications. All panels utilize color-coded synergy vectors and scalar products to quantify alignment with normative synergies.

### 2.5 Restoration of muscle synergies drives motor functional and kinematic improvements

Building on the above-described observations that synergy-based FES enhanced FM scores, joint kinematics, and synergy similarity, we next sought to uncover any correlations among the pre-to-post changes of these metrics to test the hypothesis that redeployment of the normative synergies resulting from FES-driven neuroplasticity drives clinical and kinematic recovery. Linear regression analysis revealed that improvements in FM ankle scores correlated with the degree of gait asymmetry reduction (*slope* = −0.3*, p* = 0.04, Pearson correlation coefficient), indicating that patients who achieved greater ankle symmetry demonstrated superior clinical recovery (Fig. 7a). Notably, the increase in the similarity between the stroke-affected synergies selected for FES intervention and normative synergies positively correlated with FM ankle score improvements (*p* = 0.03; Fig. 7b). Furthermore, the same metric of synergy similarity improvement correlated also with the restoration of normative knee kinematics (*p* = 0.02; Fig. 7c), as quantified by the correlation coefficients for the affected-vs-normative knee-angle trajectory comparison. With these findings, we infer that the FES-driven neural recalibration in muscle synergies drives recovery in both gait kinematics and motor functions, resulting in a more naturalistic, symmetric gait patterns and higher FM scores.

**Fig. 7.**
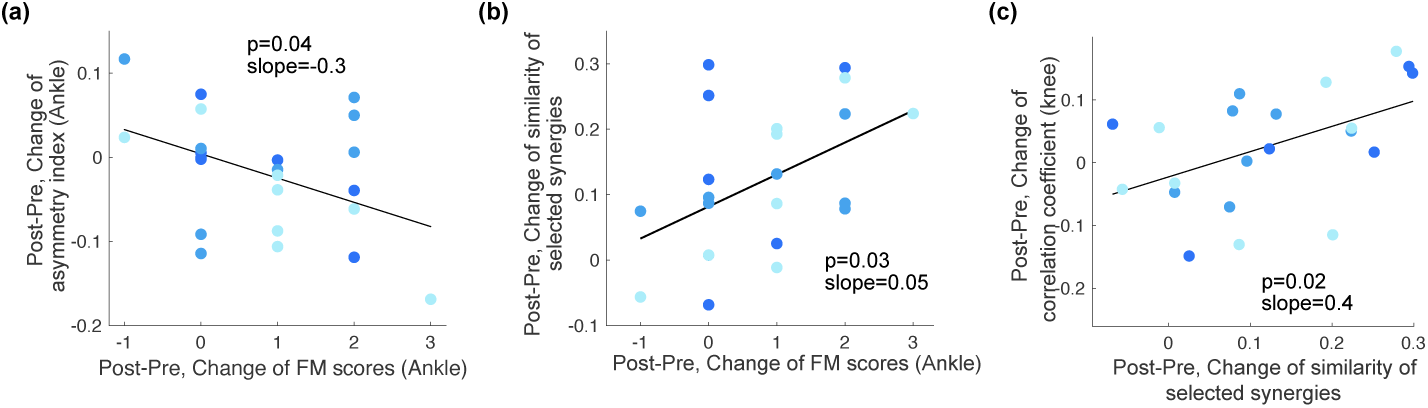
Relationships between clinical, neuromuscular, and biomechanical outcomes post-FES. (a) Linear regression between changes in FM ankle subsection scores (ΔFM) and reductions in ankle angle asymmetry (Δ asymmetry index). (b) Linear regression of ΔFM ankle scores against Δ similarity between stroke and normative muscle synergies, excluding one outlier (lowest Δ similarity). (c) Linear regression of Δ knee angle correlation to normative patterns against Δ synergy similarity, excluding one outlier (lowest Δ correlation coefficient). All analyses used Pearson’s r; significance is denoted (*p <* 0.05).

## 3 Discussion

### 3.1 Clinical improvement in chronic stroke survivors

Our five-session synergy-based FES protocol produced statistically significant improvement in the FM score for chronic stroke survivors, with sustained effects observed one month after FES. The amount of training needed for achieving a meaningful outcome here contrasts with that of conventional electrical stimulation therapies, which typically require far more treatment sessions. Previous upper limb interventions involved, for instance, 10–120 sessions [37] while lower limb interventions, 12–48 sessions [23]. Notably, prior FES efficacy research for both the upper [38] and lower [39] has largely focused on studying acute or subacute stroke populations (*<* 2 months post-stroke), with few studies demonstrating FM score improvement in chronic survivors. The significant post-FES improvement in the ankle and knee FM sub-scores observed in our FES group — absent in the sham group — suggests that task-specific synergy-based stimulation may enhance neuroplasticity even with a relatively lower dosage.

The sustained benefit observed in our cohort addresses a critical limitation of existing FES therapies. While previous studies either lack follow-up assessments [24, 39, 40] or demonstrate no long-lasting effects from the intervention [41, 42], our longitudinal analysis reveals FM score improvement that not just persisted, but even slightly increased from +2.6 immediately after FES to +2.9 at one-month follow-up (Fig. 2e). This sustainability in FM underscores the potential of synergy-based interventions to induce neurophysiological adaptations beyond temporary functional gains.

### 3.2 Gait improvements with multi-muscle FES intervention

Our findings demonstrate that a multi-channel FES system, designed to activate coordinative muscle groups in a personalized way, may significantly enhance ankle function and produce more natural ankle- and hip-joint gait patterns in chronic stroke survivors (Fig. 4). Our results are particularly noteworthy given the post-stroke prevalence of foot drop, a disabling condition affecting approximately 20% of stroke survivors, and in which weakened dorsiflexor muscles impair heel contact during walking, thus often leading to the emergence of compensatory strategies such as leg circumduction or excessive hip flexion [43, 44]. Notably, our intervention achieved measurable functional improvements even in stroke survivors with long-standing impairments (87–232 months post-stroke), suggesting the potential applicability of synergy-based FES beyond the conventional recovery window derived from observations in subacute survivors [20, 45].

While prior FES research has focused on restoring the activities of isolated muscles to address specific motor deficits (e.g., foot drop), the restoration of natural gait coordination remains understudied. For instance, single- or dual-channel systems have primarily targeted muscles such as the tibialis anterior or hamstrings to address foot drop or knee hyperextension [20, 21, 46]. Although such approaches improve dorsiflexor strength or spasticity [47], they often neglect the synergistic activation of multiple muscle groups required for inter-joint coordination, a critical factor for restoring biomechanically efficient gait. Recent advancements in multi-channel FES systems have shown promise in enhancing joint mobility during training. For example, Van et al. reported that a multi-channel FES system applied to chronic stroke survivors elicited ankle dorsiflexion and promoted knee flexion in a randomized controlled trial [45]. Similarly, Zhang et al. observed enhanced ankle and knee mobility in subacute patients using a six-channel FES protocol [48]. However, these systems remain limited by their reliance on gait-event-triggered stimulation, which activates muscles based on simplified kinematic criteria rather than neuromuscular coordination principles, thus failing to address deficits in multi-muscle coordination patterns.

In contrast, our approach integrates a principle of natural motor control with the construction of personalized FES stimulation waveform, thereby stimulating multiple muscles to mimic natural neuromotor coordination and leveraging the inherent control of the complex human neuromuscular system. Our approach yields two key advances. First, stroke survivors in the long-chronicity subgroup exhibited a significant increase in ankle dorsiflexion during overground walking as compared with the sham group after just five FES sessions (*p* = 0.01, Fig. 4a). Second, kinematic analysis reveals hip-joint trajectories in the long-chronicity subgroup aligning more closely with the normative patterns (Fig. 3, Fig. 4e). These improvements essentially translate to reduced foot drop severity and a more natural gait pattern, even in survivors with persistent deficits before FES.

### 3.3 Restoration of normative muscle synergies for functional recovery after stroke

Our study proposes that FES that targets muscle synergies — fundamental modules of neuromotor control utilized by the CNS for generating locomotion [49–52] — can promote recovery by recalibrating pathological neuromuscular coordination. While conventional assessments such as the FM score [53] and other kinematic variables [54] remain widely used for evaluating motor deficits, their limitations in precisely capturing motor impairment or predicting clinical outcomes have received growing attention [55, 56]. Some studies emphasize bio-inspired strategies that integrate interventional approaches with neurophysiological principles [28] derived from brain-imaging [57], brain signals [58] or muscle synergies [40, 59] to optimize therapeutic outcomes. Muscle synergies have been demonstrated to be reorganized through neuroplasticity during skill acquisition or recovery [60–62]. Furthermore, muscle synergies have been proposed as feasible post-stroke markers that convey information on the residual motor functions and the neurological abnormalities underlying the impairment [35, 36]. By designing FES protocols that selectively activate synergies derived from healthy patterns, we aimed here to recalibrate specific pathological neuromuscular coordination towards its expected normative patterns.

Unlike prior studies that applied the same standardized synergy-based FES pattern to all stroke survivors in the cohort [40, 59], our protocol tailored the stimulation profiles to the specific deficit of each individual at the muscle synergy level. Using EMG data from 24 age-matched healthy subjects as a normative benchmark, we identified and prioritized the impaired muscle synergies in each stroke survivor for targeted restoration by constructing the FES pattern with their corresponding normative muscle synergies. By synchronizing FES with active gait performance during the intervention, we hypothesize that the sensory feedback from FES reinforces the activation of these normative synergies within the CNS, thereby promoting neuroplastic adaptation. Our results support this hypothesis: subjects who received synergy-based FES exhibited increased similarity between their post-FES muscle synergies and the normative baseline, and this was not observed in the sham group (Fig. 5a). Most importantly, improvements in the FM score and gait-pattern correlation coefficients could be directly linked to targeted synergy modifications (Fig. 7), suggesting that restoring normative synergies enhances functional mobility. These findings align with previous studies that argue that the muscle synergies identified at a time point may serve as an outcome measure that reflects how well the subject has recovered [63, 64], and may be linked to the biomechanical parameters such as muscle strength and range of ankle motion [65, 66].

### 3.4 Limitations and future directions

This study has several limitations. First, the observed kinematic improvements exhibited significant inter-subject variability, particularly within the mid-chronicity subgroup whose gait changes were less pronounced. This heterogeneity may stem from the broad range of post-stroke chronicity in the treatment group (67.3 ± 53.1 months post-stroke), underscoring the challenge of standardizing interventions for a population with diverse recovery timelines. Second, while our synergy-driven FES protocol prioritizes personalized stimulation, the selection criteria for the normative synergies could be further refined to better address individual motor deficits. Third, the small sample size and relatively small number of FES sessions may limit the generalizability of our findings. Future studies should incorporate larger cohorts, different numbers of sessions for charting the dose-response curve, and analyses that are stratified by impairment severity or lesion location to better delineate the therapeutic efficacy in different patient subgroups.

To enhance clinical translation, subsequent work should also explore scalable implementations of synergy-based FES. For instance, integrating wearable sensors with closed-loop FES systems could enable real-time synergy monitoring and adaptive stimulation in home-based settings. Additionally, investigating the neurophysiological mechanisms underlying synergy restoration could refine therapeutic targeting and deepen our understanding of motor recovery in chronic stroke.

## 4 Methods

### 4.1 Subjects

We recruited 33 community-dwelling chronic stroke survivors with post-stroke duration from 8 to 232 months. Subjects were assigned to either the FES group (*N* = 23; 13 females; age = 58.9 ± 8.2) or the sham group (*N* = 10; 5 females; age = 63.2 ± 5.3 years old). Those in the former group received genuine FES intervention while those in the latter received sham FES without stimulation. All subjects had sustained brain damage due to a cerebrovascular accident. Other detailed information about the participants is shown in Table 1. All procedures were approved by The Joint Chinese University of Hong Kong - New Territories East Cluster Clinical Research Ethics Committee (CUHK-NTEC CREC) (protocol no. 2019.498). All subjects gave informed consent before experimentation. The study design complies with all relevant regulations regarding human experimentations. This study protocol was also registered at ClinicalTrials.gov (protocol no.NCT04155866 and no.NCT04154514).

**Table 1.** Participants demographics.

| Demographic variables | FES group (n=23) | Sham group (n=10) | p-values |
| --- | --- | --- | --- |
| Age (year) | 58.9±8.1 | 63.2±5.3 | p=0.14 |
| Sex (male/female) | 10/13 | 5/5 | p=1.0 |
| Type of lesion (ischemic/hemorrhagic) | 10/11 | 5/5 | p=1.0 |
| Side of hemiplegia (right/left) | 8/15 | 6/4 | p=0.26 |
| Stroke chronicity (month) | 67.3±53.1 | 98.1±90 | p=0.33 |
| Height (cm) | 162.0±7.7 | 163.7±8.3 | p=0.48 |

### 4.2 Experimental procedure

In our study, participants underwent five sessions of genuine or sham personalized FES. Each session consisted of more than 600 steps of overground walking while electrical stimulations were delivered to 6−7 muscles on the stroke-affected lower limb. To ensure safety, some of the subjects with more severe motor impairment walked with a movable harness (Lite-Gait; Mobility Research, Temple, AZ, USA) during FES intervention to prevent accidental falls. All intervention sessions were completed within 20 ± 11 days for all stroke survivors.

Prior to the intervention, we conducted a pre-FES assessment, which involved recording lower-limb kinematics and bilateral electromyographic signals (EMG) of lower limb muscles while participants walked overground at their self-selected speed. Additionally, the Fugl-Meyer Assessment of Lower Extremity (FM) was conducted. These recorded data were then used to design personalized stimulation patterns. After completing all five intervention sessions, a post-FES assessment was performed immediately for all participants. One month after the intervention, a follow-up assessment was conducted to determine whether the improvements achieved through FES were sustained. The experimental procedures are illustrated in Fig. 1.

### 4.3 Data recordings

In each assessment, bilateral kinematic data were collected from the lower limbs and trunk of each patient during overground walking using a 3-dimensional camera motion capture camera system (Vicon MX; Oxford Metrics Group, Oxford, UK) at a sampling rate of 100 Hz. Infrared reflective markers (*N* = 35) were attached to each participant according to the lower-body plug-in gait model [67, 68] to track limb movement.

To assess muscle activities during overground walking, wireless electrodes (Trigno, Delsys; Natick, MA, USA) were attached to the skin surface to record EMG signals at 2000 Hz. EMG recordings were obtained from 14 trunk and lower limb muscles on each side, including tibialis anterior (TA), medial (MG) and lateral heads of the gastrocnemius (LG), soleus (Sol), vastus lateralis (VL), vastus medialis (VM), rectus femoris (RF), hamstrings (Hams), adductor longus (AL), tensor fascia latae (TFL), gluteus maximus (GM), erector spinae (ES), external oblique (ExtO) and latissimus dorsi (Lat-Dor). The placement positions of the electrodes were identified using guidelines from the Non-Invasive Assessment of Muscles-European Community Project (SENIAM). Before EMG sensor attachment, the skin over the attachment positions was cleaned by alcohol (75%). Then the sensors were attached to the skin surface with double-sided tape, and stabilized in position with self-adherent bandage wrap (3M Coban^TM^). Finally, lower-extremity FM assessment was performed for each stroke survivor.

### 4.4 Kinematics analysis

After data collection, the kinematic data were analyzed using Visual3D (V3D; C-Motion, Washington DC, USA). By utilizing the spatial trajectories of the Vicon markers, this software calculates the 3-dimensional ankle, knee, and hip angles based on the segment coordinate systems in V3D. In this study, we focused on analyzing joint angles in the sagittal plane, and in particular, the degree of extension and flexion of each joint. All joint angle profiles were processed through a low-pass Butterworth filter set at 6 Hz. Gait cycles were identified by using times of heel strikes, defined by representing the coordinates of the heel and toe markers with the pelvis coordinate system [69]. Gait cycles with high data quality were selected for further analysis. Finally, all gait cycles from the participants were resampled to 200 time points for subsequent analyses.

### 4.5 EMG pre-processing

The EMGs collected from the both sides of each subject were preprocessed with the following steps: removal of noise from powerline interference [70, 71], high-pass filtering (cutoff at 40 Hz), rectification, low-pass filtering (40 Hz), integration (over 20-ms intervals) and variance normalization of each muscle [72]. The cutoff frequencies of the high- and low-pass filters were selected to preserve the most information in the EMGs while keeping the number of extracted synergies the same even when higher or lower cutoff frequencies were used for the high- or low-pass filters, respectively [73]. All EMGs were analyzed using custom functions written in Matlab (Mathworks; Natick, MA, USA).

### 4.6 Muscle synergy identification

Muscle synergies were extracted from the multi-channel EMG signals using the nonnegative matrix factorization algorithm (NNMF) [31]. Let **D** be a non-negative *m* × *n* data matrix comprising *n* samples of an *m*-dimension data vector. The NNMF models **D** to be a linear combination of time-invariant synergy vectors **W** and time-varying activation coefficients **C**, such that

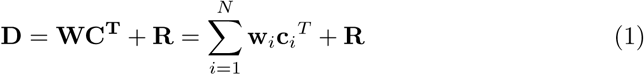

where vector **w***_i_* is the *i^th^* column of **W** denoting the *i^th^* muscle synergy, and vector **c***_i_*, *i^th^* column of **C**, is the temporal activation coefficients for **w***_i_*. **R** is the residual unexplained by the model.

To identify the number of muscle synergies needed to reconstruct the EMGs adequately, we successively increased the number of synergies extracted from one to the number of muscles recorded and selected the minimum number of synergies required for an EMG-reconstruction *R*^2^ of 80% [62]. The *R*^2^ was calculated as follows,

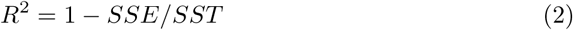

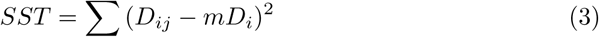

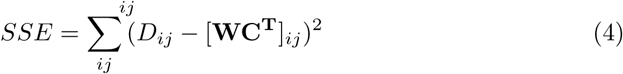

where SST is the sum squared total, *D_ij_* is the EMG data of the *i*th muscle at the *j*th time point, *mD_ij_* is the average EMG value of the *i*th muscle, and SSE is the sum squared error. To prevent the extracted synergy set from representing a suboptimal solution located at a local minimum on the surface of EMG reconstruction error, at each number of synergies, NNMF was run 20 times, each time with different initial estimates of **W** and **C** whose matrix components were uniformly distributed between 0 and the maximum EMG amplitude. The run with the highest *R*^2^ was selected for further analyses.

### 4.7 Design of personalized synergy-based FES stimulation pattern

To design a stimulation pattern that guides the impaired muscle synergies on the stroke-affected leg to change towards the normative patterns, a set of normative muscle synergies is obviously needed to serve as a baseline reference. Here, to obtain this baseline, we utilized a database of lower-limb EMG that we previously described [73]. This database contains EMGs of the same 14 muscles obtained from a group of age-matched neurologically intact subjects (*N* = 24) during overground walking. After conducting a pre-FES assessment for each stroke survivor, we selected one normal subject from this normative database using the cross-fit method [74]. This method involves using the normative synergy set from each normal subject in the database to reconstruct the stroke-affected-side EMG activities of the stroke survivor. The normal subject whose synergies yielded the highest cross-fit *R*^2^ value was selected as the subject best suited to provide the normative synergy set for that stroke survivor.

Subsequently, each normative synergy of the selected normal subject was matched to a stroke-affected-side synergy of the stroke survivor by maximizing the synergies’ pairwise similarity. Similarity between the muscle synergies of the two subjects was quantified using the scalar product after the synergies were normalized to unit vectors. The normative synergies that were poorly matched to the stroke-affected-side synergies, as indicated by their low similarity values, are also the ones missing in the synergy repertoire of the stroke survivor; they are therefore also the synergies that should ideally be restored in the stroke survivor. Here, we set the dissimilarity scalar-product threshold to be 0.8 (the threshold of two stroke patients was set at 0.85), so that the normative synergies with a similarity lower than 0.8 were selected for the design of the FES stimulation pattern. As illustrated in Fig. 1, the personalized FES waveform was constructed by multiplying all the selected normative muscle synergies by their corresponding normative temporal activations as indicated in the data of the selected normal subject for that stroke survivor. Critically, the stimulation waveform was based purely on muscle synergy deficit, without regard to the kinematics produced by the superposition of the FES waveforms and the motor commands produced by the subject’s motor system. Subsequently, the stance and swing portions of the constructed FES waveform for each gait cycle were temporally rescaled to align with the natural walking cadence and stance ratio of the stroke survivors. Finally, the stimulation amplitude for each muscle was adjusted through a sequential process: initially scaling each muscle’s normalized partial EMG pattern by its corresponding maximum tolerable current of the patient, followed by multiplying the resultant value by the variance ratio between the partial and its full normative EMG signals derived from the healthy subject. The pulse width and frequency of the stimulating waveforms were set at 200 *µ*s and 50 Hz, respectively [75]. The pulse amplitude ranged from 0 to 30 mA in our study.

### 4.8 Statistics

In this study, two-tailed unpaired t-tests were employed to compare differences between the means of different groups while two-tailed paired t-tests were used to assess longitudinal changes of means within the same group. Homogeneity of variance was assessed as a prerequisite for conducting the unpaired t-tests. Pearson’s correlation coefficient was applied to evaluate the significance of associations between variables, and linear regression was utilized to model relationships between paired samples. The significance threshold for hypothesis testing was set at *α* = 0.05. All statistical analyses were performed in MATLAB.

## 5 Conclusion

In this study, we developed a personalized, synergy-based FES intervention for lowerlimb rehabilitation in chronic stroke survivors, where muscle synergies served as a biomarker to reflect motor deficits in subjects with cortical lesions during movements. After five sessions of intervention, participants receiving genuine FES treatment demonstrated superior clinical recovery and more natural gait patterns compared to the control group. Critically, the intervention selectively enhanced the similarity between stroke-affected muscle synergies targeted by FES and their normative synergies used in FES design. Notably, this improvement correlated with the gains in lower-limb FM score and the enhancement in gait symmetry. These findings validate our hypothesis that multi-channel FES grounded in muscle synergies can guide residual neural circuits toward normalized activation patterns by leveraging neuroplasticity to recruit normative synergies. This approach advances targeted neuromodulation strategies for restoring gait control modules in stroke rehabilitation.

## Data Availability

The datasets in the present study are not publicly available.

## Acknowledgments

This work was funded by the Research Grants Council of the Hong Kong Special Administrative Region, China (Project Nos. R4022–18, N CUHK456/21, 14114721, 14119022, and STG1 M-401 24-N to V.C.K.C.), grants from The Chinese University of Hong Kong (Project Nos. 2020095, 2021.065 for Faculty of Medicine, and project ”Impact Case C7” for Research Committee, to V.C.K.C.).

## Declarations

The co-authors declare that the research was conducted in the absence of any commercial or financial relationships that could be construed as a potential conflict of interest.

